# Attributing non-specific symptoms to cancer in general practice: a scoping review

**DOI:** 10.1101/2025.03.27.25324367

**Authors:** Gladys Langue, India Pinker, Valerie Moran, Sophie Pilleron

## Abstract

**Background:** Non-specific cancer symptoms are challenging to interpret in general practice. They can be attributed to a wide range of other conditions and delay the cancer diagnosis, increasing the risk of poor outcomes.

**Aim:** To summarise existing knowledge on the attribution of non-specific symptoms to potential cancer in general practice and identify gaps in the literature.

**Method:** We conducted a scoping review, following Joanna Briggs Institute’s guidance and reported according to the PRISMA for scoping reviews checklist. Non-specific symptoms were defined based on NICE guidelines for suspected cancer. We systematically searched six databases and search engines for original papers, systematic reviews and doctoral theses. Two reviewers independently screened titles, abstracts, full-texts and reference lists. Included articles were then uploaded to the AI-based tool ResearchRabbit to identify further papers. Findings were synthesised using the Refined Andersen Model of Total Patient Delay.

**Results:** Eight studies were included. These addressed fatigue, recurrent infection, pallor, weight loss, and deep vein thrombosis, with general practitioners (GPs) defining the latter two as cancer-specific. Factors influencing attribution to cancer included pre-existing conditions, the number, type and combination of symptoms as well as GPs’ gut feeling and knowledge, patients’ concern, and the frequency of medical visits.

**Conclusion:** This scoping review identified a limited number of studies on the attribution of non-specific symptoms to potential cancer in general practice, with symptoms such as pruritus and new-onset diabetes not addressed. It highlights the need for further research into GPs diagnostic reasoning related to non-specific cancer symptoms, especially on under-researched symptoms.

## Introduction

General practitioners (GPs) are often the entry point into the healthcare system. They play a critical role in evaluating signs and symptoms and referring patients for further investigations when a serious disease, such as cancer, is suspected [1].

Despite significant advances in diagnosis and treatment, cancer is amongst the most frequently missed diagnoses [2–4], and remains a global health challenge, with over 20 million new cases and approximately 10 million deaths estimated in 2022 [5]. Early cancer detection is essential but often complicated by the wide range of symptoms cancer can present. These range from specific manifestations, such as postmenopausal vaginal bleeding or new-onset seizures in individuals with no history of epilepsy, to non-specific symptoms (NSS), including fatigue, unintentional weight loss, and appetite loss among others [6,7].

Identifying cancer in the context of NSS is particularly challenging for GPs, even though these symptoms are present in 22% of cancer cases [8]. NSS are frequently associated with a variety of non-cancerous conditions, especially in patients with other conditions like heart failure, chronic obstructive pulmonary disease, or depression, which share symptoms such as loss of appetite and fatigability [3,9–12]. Moreover, some NSS are common in general practice, with fatigue reported by one in four patients [13]. This overlap can delay cancer diagnosis [3,14], resulting in more advanced stages at diagnosis, poorer clinical outcomes and reduced quality of life [12,15–17].

Interestingly, many cancer diagnoses are preceded by an increase in the number of general practice consultations in the months prior, suggesting missed opportunities for earlier detection at the GP level [18– 20]. However, the process by which GPs attribute NSS to potential cancer remains underexplored [21,22]. This scoping review aims to summarise existing literature on the attribution of NSS to cancer in general practice, identify influencing factors, and highlight gaps in the current literature.

## Method

The protocol is available on Open Science Framework (OSF) [23]. This scoping review was prepared using the Preferred Reporting Items for Systematic reviews and Meta-Analyses for scoping reviews (PRISMA-ScR) checklist (text S1) and Joanna Briggs Institute’s guideline [24].

### Classification of non-specific symptoms

There is no consensus on a classification of NSS in cancer. To standardise our approach, we have adopted the classification of the National Institute for Health and Care Excellence (NICE) [25]. According to the classification, NSS associated with cancer include fatigue, unexplained weight loss, appetite loss, early satiety, deep vein thrombosis, pallor, new-onset diabetes, night sweats, unexplained fever, persistent or recurrent unexplained infections, and pruritus.

### Literature review

In March 2024, we conducted a comprehensive search for peer-reviewed articles in Medline, Cumulative Index to Nursing and Allied Health Literature (CINAHL), PsycInfo and Embase databases. The search strategy including terms referring to “cancer”, “general practitioner” and “symptom attribution” was reviewed by two librarians (Tables S1-S5 and Text S2). Following the librarian’s recommendations, we excluded NSS from the search strategy, as pilot testing indicated that these symptoms frequently appear only in the full-text rather than in the title or abstract.

We included original qualitative and quantitative research articles and systematic literature reviews. There was no restriction on the period, geography and language. In July 2024, we also searched Google Scholar and Open Access Theses and Dissertations for doctoral theses published since 2022. This time range was chosen to capture recent scientific work that had not yet been published.

After the removal of duplicates, two independent reviewers screened titles, abstracts, full-texts and the reference lists of the included papers based on predefined eligibility criteria (Table 1). Any disagreements were resolved by a third reviewer. Finally, included articles in the scoping review were uploaded to ResearchRabbit, a publication discovery tool that uses artificial intelligence to identify additional references [27]. This approach aimed to enhance the comprehensiveness of the search. Fig 1 shows the selection process.

**Table 1.** Eligibility criteria for the selection of source of evidence.

| Component | Inclusion criteria | Exclusion criteria |
| --- | --- | --- |
| Population | General practitioners (GP) | Specialist medical doctors<br>Other health professionals (e.g., nurses) |
| Concept | Non-specific symptoms as defined by NICE<br>Symptom attribution by GPs<br>Suspicion of cancer<br>Justification of cancer suspicion for referral | Symptoms not categorized as non-specific by NICE<br>Symptom attribution by patients<br>Referral for reason other than suspected cancer |
| Context | Consultation at a primary care level | GP consultation within an emergency ward or other level of care |
| Study design | Any quantitative, qualitative or mixed-methods design<br>Systematic reviews | Case report<br>Narrative review |
| Publication type | Peer-reviewed articles<br>Doctoral theses from 2022 onward. The rationale for including theses from this period is to capture scientific research results that have not yet been published. | Conference abstracts<br>Editorials<br>Commentaries<br>Letters to editor |
| Language | No restriction (the authors' institution hosts diverse language expertise) | None |

**Fig 1. PRISMA flow diagram**

For each included document, we extracted first author name, year of publication, study objective(s), country(ies) where the study was conducted, type of research, study design, data collection tool, number of GPs and patients and their main characteristics (e.g., sex, years of experience), type of cancer, NSS, factor(s) influencing the symptom attribution and presentation of the factor(s). Data extraction was conducted independently by two reviewers, with discrepancies resolved through discussion between the two reviewers and, if necessary, discussion with a third reviewer. This deviates from the original protocol [23] but was implemented to optimise time and minimise the workload for the reviewers.

Results are summarised using a narrative approach. Factors influencing symptom attribution were categorised according to the Refined Andersen Model of Total Patient Delay [28]. This conceptual framework categorises the contributing factors to patient care delays (including diagnosis) into disease-(e.g., tumour site), patient-(e.g., demographic), healthcare provider-(e.g., previous experiences) and healthcare system (e.g., healthcare policy)-related factors.

We used EndNote 21.1 to manage references [29] and Rayyan for screening [30].

## Results

Out of 24,397 references, we included eight papers. The list of excluded full-texts with reasons for exclusion is available in Tables S6 and S7.

No additional articles were found using ResearchRabbit. However, for unknown reasons, it was possible to export only 3,843 of the 3,844 identified references and we could not identify the missing reference.

### Characteristics of included studies (Table 2)

Of the eight included articles [31–38], there were four qualitative studies [31–34], two quantitative studies [35,36], one mixed-method study [37], and one systematic review [38]. Most studies were conducted in Europe [31–36,38], with one study originating from the United States [37]. GPs’ years of practice in the included samples ranged from two to 44 years [31–34,36,37].

**Table 2.**
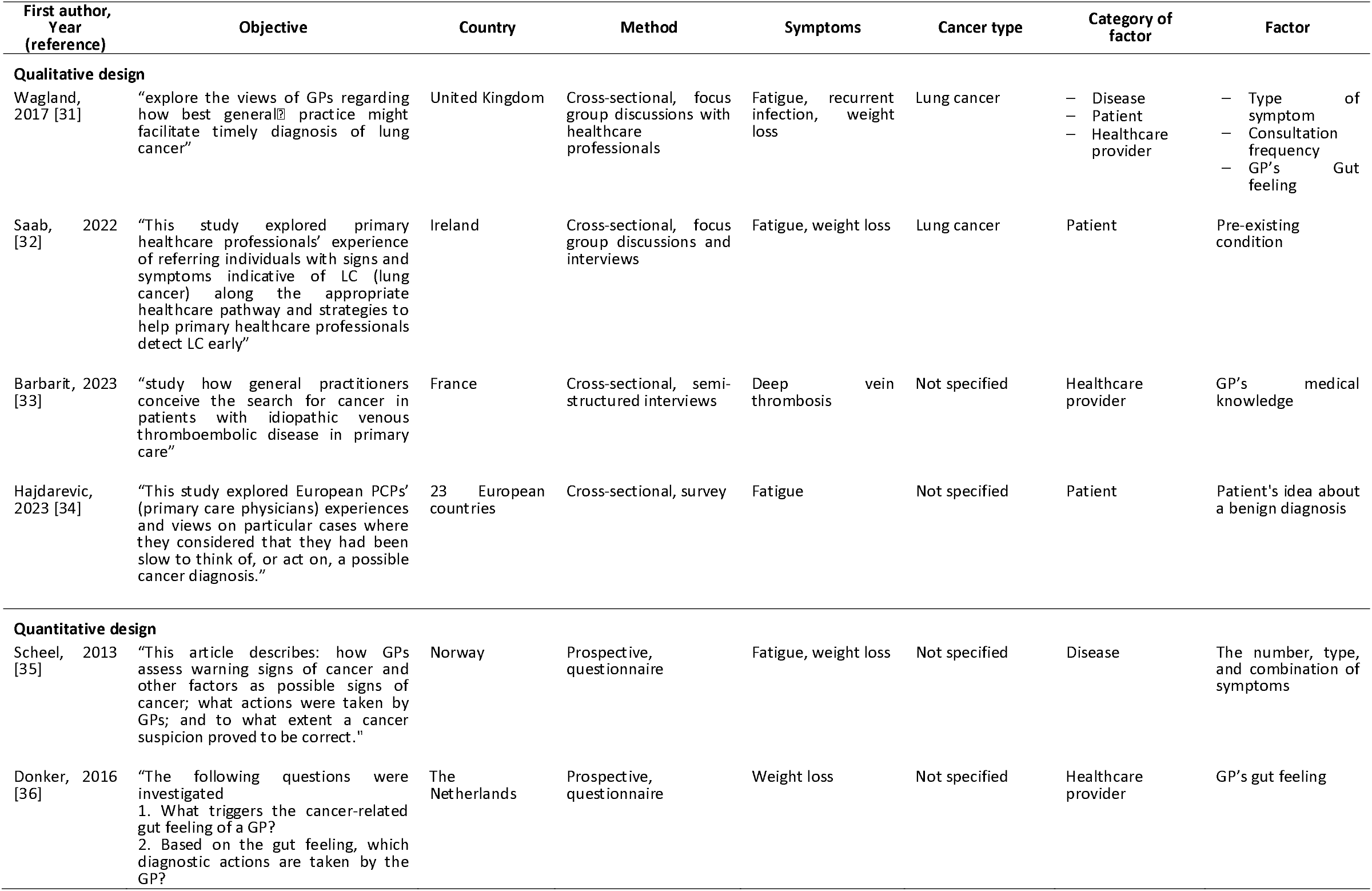
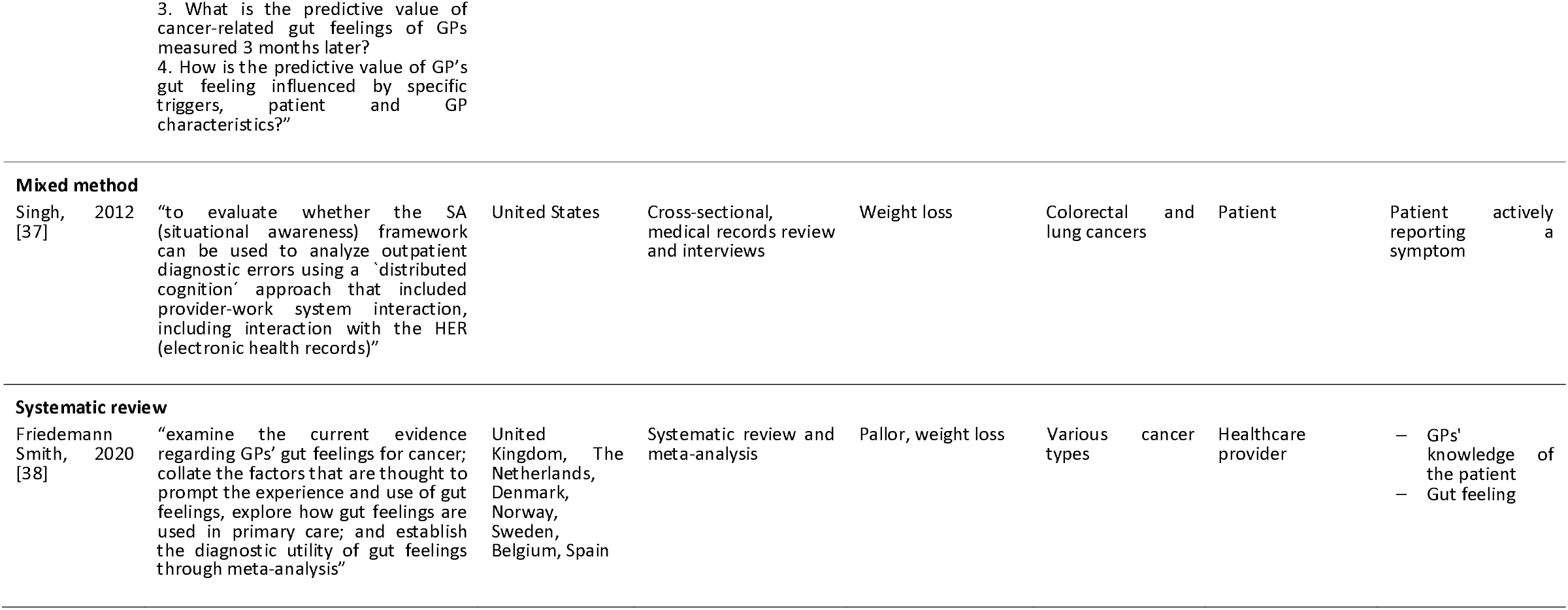
Characteristics of included studies.

None of included papers explicitly focused on the attribution of NSS to cancer in general practice. While the study by Barbarit and colleagues specifically focused on deep vein thrombosis (18), the remaining seven studies mentioned NSS without making them the primary focus of their investigation.

Weight loss was the most frequently cited NSS [31,32,35–38], followed by fatigue [31,32,34,35]. Recurrent infection and pallor were mentioned in two studies [31,38]. We did not identify any studies mentioning or discussing factors influencing the attribution of new-onset diabetes, pruritus, night sweats, appetite loss, early satiety or unexplained fever to a potential cancer.

A limited number of factors influencing GPs’ symptom attribution were identified, encompassing disease-, patient- and healthcare provider-related factors.

### Disease factors

Not all NSS are regarded equally in the context of cancer suspicion. Despite being classified as NSS, three studies reported that weight loss [31,35] and deep venous thrombosis – particularly when of unclear origin – [33] quickly raised concerns about a potential cancer, even when they occurred in isolation. In contrast, fatigue generally did not elicit concern from GPs unless it was accompanied by other symptoms [31]. In the study conducted by Scheel and colleagues, cancer suspicions were higher with an increasing number of symptoms: GPS suspected cancer in 68 out of 662 (10.3%) fatigue registrations in medical records when fatigue was the sole symptom, whereas the registration of fatigue in combination with at least two warning signs of cancer elicited a suspicion of cancer in 76 out of 137 (55.5%) cases [35].

However, the specific combination of symptoms was also important. Bleeding and fatigue showed a positive association with cancer suspicion (OR = 6.5, 95% Confidence Interval (CI) 1.7 to 24.7) but the combination of fatigue and pain (OR = 0.4, 95% CI 0.2 to 0.7) or that of digestive problem, fatigue, and pain (OR = 0.3, 95% CI 0.0 to 0.8) showed a negative association [35].

### Patient factors

Three studies identified patient-related factors that prompt GPs to suspect cancer [31,32,37] and one highlighted a factor that can deflect suspicion [34].

The way patients communicate their concerns can elicit a suspicion of cancer. In the study of Singh and colleagues, one GP noted that despite documenting the patient’s weight loss in the medical record, he did not recognise its significance until the patient explicitly expressed concern [37].

Conversely, when a patient attributes their symptoms to a benign condition, it may discourage the GP from pursuing further investigation. This was illustrated in the study of Hajdarevic and colleagues, where a GP recounted:

> An employee around 60 years at my former clinic came to me with a fatigue. She had been feeling tired from [for] half a year. She had been able to change her work content avoiding activities she felt increased the fatigue. […] Both I as a doctor and she as a patient zoomed in on “burn-out”. [34, p7]

The study of Wagland and colleagues revealed that the frequency of patient consultations may also influence the suspicion of cancer [31]. The authors reported that a patient who rarely visits a GP tends to prompt greater concern from the GP, regardless of the reported symptoms, compared to a patient who consults regularly.

Pre-existing conditions may also contribute to delayed cancer diagnoses, as highlighted by Saab and colleagues [32]. In this study, one GP described attributing a patient’s weight loss to chronic obstructive pulmonary disease rather than considering it as a potential symptom of cancer.

### Healthcare provider factors

The attribution of NSS can also be influenced by medical training, as suggested by the study conducted by Barbarit and colleagues [33]. In this study, GPs acknowledged that their diagnostic reasoning was shaped by their training. Specifically, they reported that an atypical presentation of deep venous thrombosis would automatically raise suspicion of cancer, as this association had been emphasised during their medical education.

GPs familiarity with the patient was also important. The study by Friedemann Smith and colleagues showed that GPs who were well-acquainted with their patients were better positioned to detect changes in appearance, such as alterations in skin tone or weight loss [38].

Three studies examined GPs’ gut feeling [31,36,38]. A gut feeling, when raising concern, was defined as “an uneasy feeling perceived by a GP as he/she is concerned about a possible adverse outcome, even though specific indications are lacking” [39]. It arose particularly in patients presenting with weight loss and a change in their appearance. Although various types of symptoms can trigger gut feelings, interviewed GPs perceive it as particularly valuable when assessing NSS [31].

## Discussion

To our knowledge, this is the first scoping review to explore the attribution of NSS to a potential cancer by GPs. Despite the recognised challenge of NSS in general practice, our scoping review retrieved only eight studies, which suggests that this topic has received limited research attention.

The inherent difficulty in studying diagnostic reasoning may explain the limited number of studies included in our scoping review. Symptom attribution is a critical component of this reasoning and occurs within a GP’s mind. Although some GPs record aspects of their diagnostic reasoning in patient health records, this practice is not standardised and varies widely across practitioners [40,41]. Additionally, GPs may not be aware of all factors influencing their diagnostic reasoning [21]. This makes the systematic study of this cognitive process particularly challenging.

We identified a small number of factors that may influence the suspicion of cancer when a patient presents with NSS. Some factors identified as raising cancer suspicions included specific symptom types or combinations [33,35], patients who rarely consult a GP [31] and the GPs’ gut feeling [31,36,38]. A previous systematic review on cancer-related gut feeling further supports the use of gut feeling as a legitimate criterion for suspected cancer referral [42].

In contrast, pre-existing conditions [32] or the attribution of symptoms to another illness by the patients [34] may lead to cancer not being considered. This highlights the extent to which NSS of cancer can easily be attributed to other conditions. Although factors such as patient sex, history of cancer, symptom duration, GP’s age and patients risk-taking attitude have been identified in the literature as influencing symptom attribution to cancer [35,43], these were not identified in our scoping review. It may be due to our exclusive focus on NSS.

Some NSS listed by NICE are uncommon features of cancer [6,33]. For example, night sweats have a maximum reported prevalence of 6% in patients with Hodgkin lymphoma, while new-onset diabetes is seen only in 1% of pancreatic cancer cases [6]. Due to their low prevalence, these symptoms may be given less priority in research and therefore, be under-recognised by GPs as a potential cancer symptom.

Of the eleven NSS listed by NICE [25], data were available for only five, namely weight loss, fatigue, recurrent infection, pallor and deep venous thrombosis. Among these, weight loss is consistently identified as an alarm symptom, rapidly prompting GPs to consider cancer. This suggests that it may be inappropriate to include weight loss in the list of NSS. This was corroborated during formal discussions with GPs as part of the Public and Patient Involvement activities conducted within the framework of preparing the “Reducing Disparities in Cancer Outcomes – Qualitative component” research project [44]. A deep venous thrombosis, particularly when occurring in atypical contexts, also quickly elicits the suspicion of cancer.

Although included studies have been conducted in different countries, none have provided information on the potential association between the type of healthcare system and the association between NSS and a potential cancer. Consequently, we do not know whether a healthcare system that implements a cancer diagnostic pathway for NSS, such as those in Denmark or United Kingdom [22], facilitates earlier consideration of cancer by GPs compared to a system without these pathways. The existence of these pathways may signify a broader recognition of NSS as being potentially indicative of cancer, thereby encouraging their consideration in clinical practice.

Finally, despite evidence that comorbidity and multimorbidity can delay cancer diagnosis [12,45–47], few studies have investigated GPs’ diagnostic reasoning processes in the context of multimorbidity and NSS. Our scoping review identified only one study [32] addressing this issue, yet it offers limited insights, noting only that pre-existing conditions tend to reduce the likelihood of GPs suspecting cancer [32]. However, given population ageing and the rise in the incidence of chronic diseases and cancer [48,49], GPs will increasingly treat multimorbid patients who present with NSS. Understanding how GPs consider the possibility of cancer in these complex cases is essential to support medical doctors in the early detection of cancer.

### Strengths and limitations

The strengths of this scoping review include the absence of restrictions on language and time period, and the search for grey literature. Additionally, we maximised our chances to retrieve relevant papers by utilising ResearchRabbit. Limitations include the lack of standardisation and consistent definition of the concept of “symptom attribution”, which may have reduced the specificity of the chosen keywords. Different classifications of NSS exist in the wider scientific literature, some of which including symptoms beyond those listed by NICE, such as abdominal or back pain, bruising, nausea or vomiting [6,8,50,51]. While NICE’s classification benefits from being developed through expert consensus and provides a clear, structured framework, the origins of other classifications remain unclear. Consequently, papers using NSS symptoms not included in the NICE classification were excluded, potentially excluding relevant literature.

## Conclusion

This scoping review identified a limited number of studies on the attribution of NSS to potential cancer in general practice. It highlights the need for further research into this topic, particularly as the older population and the number of people living with chronic diseases continues to grow. Future studies should prioritise investigation into underexplored NSS, including new-onset diabetes, pruritus, night sweats, loss of appetite, early satiety and unexplained fever.

## Data Availability

All relevant data are within the manuscript and its Supporting Information files.

## Acknowledgements

The authors would like to thank Coralie Dessenne, a librarian at the Luxembourg Institute of Health, and Dr Claire Godet, a specialist librarian at the Luxembourg Learning Centre, for their help in developing the search strategy. The authors acknowledge the use of ChatGPT-3.5 to enhance the clarity of the manuscript.

## Supporting information

S1 Text. PRISMA for scoping reviews

S1 Table. Search strategy for Cumulative Index to Nursing and Allied Health Literature (CINHAL) (26/03/2024) S2 Table. Search strategy for Medline via PubMed (26/03/2024)

S3 Table. Search strategy for PsycInfo via Ovid (26/03/2024) S4 Table. Search strategy for Embase (26/03/2024)

S2 Text. Search strategy for Google Scholar

S5 Table. Search strategy for Open Access Theses and Dissertation (11/07/2024)

S6 Table. Excluded articles with reason for exclusion, ordered by year of publication and first author name

S7 Table. Articles retrieved by ResearchRabbit we excluded with reasons for exclusion, ordered by year of publication and first author name

