## Supplemental Table 1 for "Attributing non-specific symptoms to cancer in general practice: a scoping review"

### S1 Table. Search strategy for Cumulative Index to Nursing and Allied Health Literature (CINHAL) (26/03/2024)

| **#** | **Query** | **Limiters/Expanders** | **Results** |
| --- | --- | --- | --- |
| S1 | (MM "Neoplasms+") |  | 590,712 |
| S2 | TI (adenocarcinoma* or blastoma* or cancer* or carcinoma* or glioblastoma* or hepatoblastoma* or hodgkin* or leuk#emia* or lymphoma* or malignan* or medulloblastoma* or melanoma* or myeloma* or neoplas* or nephroblastoma* or nonhodgkin* or "non-hodgkin*" or onco* or pancreatoblastoma* or retinoblastoma* or sarcoma* or tumo#r*) |  | 628,684 |
| S3 | S1 OR S2 |  | 797,668 |
| S4 | MH "Family Practice" |  | 26,631 |
| S5 | MH "Physicians, Family" |  | 23,66 |
| S6 | TI ((family or general) n1(doctor* or medicine or physician* or practi*)) |  | 21,52 |
| S7 | TI (GP or GPs) |  | 6,503 |
| S8 | TI (generalist*) |  | 654 |
| S9 | MH "Primary Health Care" |  | 74,551 |
| S10 | TI ("primary n2 care") |  | 0 |
| S11 | TI ("primary n2 care") |  | 890 |
| S12 | TI ("primary care") |  | 38,327 |
| S13 | S4 OR S5 OR S6 OR S7 OR S8 OR S9 OR S10 OR S11 OR S12 |  | 131,275 |
| S14 | (MH "Clinical Reasoning") |  | 985 |
| S15 | TI "clinical reasoning" OR AB "clinical reasoning" |  | 3,475 |
| S16 | (MH "Patient Assessment+") |  | 77,24 |
| S17 | TI (assess*) OR AB (assess*) |  | 1,076,016 |
| S18 | (MH "Causal Attribution") |  | 7,625 |
| S19 | TI (attribut*) OR AB (attribut*) |  | 77,471 |
| S20 | (MH "Decision Making, Clinical+") |  | 38,304 |
| S21 | TI ("clinical decision making") OR AB ("clinical decision making") |  | 10,316 |
| S22 | TI "first impression" OR AB "first impression" |  | 217 |
| S23 | (MH "Perception+") |  | 95,377 |
| S24 | TI perception OR AB perception |  | 155,788 |
| S25 | TI recogni* OR AB recogni* |  | 165,062 |
| S26 | TI (suspicion* OR suspicious OR suspect*) OR AB (suspicion* OR suspicious OR suspect*) |  | 69,481 |
| S27 | TI apprais* OR AB apprais* |  | 31,092 |
| S28 | TI (think OR thinks OR thought OR thinking) OR AB (think OR thinks OR thought OR thinking) |  | 100,227 |
| S29 | (MH "Referral and Consultation+") |  | 59,562 |
| S30 | TI (refer* NOT (reference* OR referencing)) OR AB (refer* NOT (reference* OR referencing)) |  | 139,752 |
| S31 | TI referral* OR AB referral* |  | 60,081 |
| S32 | TI ((refer n2 patient) OR (refers n2 patient) OR (refer n2 patients) OR (refers n2 patients)) OR AB ((refer n2 patient) OR (refers n2 patient) OR (refer n2 patients) OR (refers n2 patients)) |  | 2,39 |
| S33 | S14 OR S15 OR S16 OR S17 OR S18 OR S19 OR S20 OR S21 OR S22 OR S23 OR S24 OR S25 OR S26 OR S27 OR S28 OR S29 OR S30 OR S31 OR S32 |  | 1,743,043 |

**S1 Table** (continued)

| **#** | **Query** | **Limiters/Expanders** | **Results** |
| --- | --- | --- | --- |
| S34 | S3 AND S13 AND S33 |  | 2,076 |
| S35 | S3 AND S13 AND S33 | Limiters - Publication Type: Case Study | 53 |
| S36 | S34 NOT S35 |  | 2,023 |
