## Supplemental Text 2 for "Attributing non-specific symptoms to cancer in general practice: a scoping review"

### S2 Text. Search strategy for Google Scholar

11/07/2024

2022- 2024: allintitle: general practitioner AND cancer diagnosis (4 results)

2022 - 2024: allintitle: general practice AND cancer diagnosis (5 results)

12/07/2024

2022 - 2024: allintitle: general practitioner AND cancer AND diagnose (0 result)

2022 - 2024: allintitle: GP AND cancer AND diagnosing (0 result)
