## Supplemental Table 6 for "Attributing non-specific symptoms to cancer in general practice: a scoping review"

### S6 Table. Excluded articles with reason for exclusion, ordered by year of publication and first author name

| **First author, date** | **Title** | **Reason for exclusion** |
| --- | --- | --- |
| Fetter, 1952 | Recognition of urological malignancies by the general practitioner | Wrong publication type |
| Nylenna, 1986 | Diagnosing cancer in general practice: when is cancer suspected? | No focus on non-specific symptoms |
| Nylenna, 1986 | Diagnosing cancer in general practice: from suspicion to certainty | No focus on non-specific symptoms |
| Burgess, 1998 | Who and what influences delayed presentation in breast cancer? | No focus on non-specific symptoms |
| Burgess, 2001 | A qualitative study of delay among women reporting symptoms of breast cancer | No info on diagnostic reasoning |
| de Nooijer, 2001 | A qualitative study on detecting cancer symptoms and seeking medical help; an application of Andersen's model of total patient delay | No info on diagnostic reasoning |
| Mariscal, 2001 | Determinants of the interval between the onset of symptoms and diagnosis in patients with digestive tract cancers | No focus on non-specific symptoms |
| Davies, 2002 | Cancer recognition and primary care | Wrong publication type |
| Jiwa, 2004 | Less haste more speed: Factors that prolong the interval form presentation to diagnosis in some cancers | No focus on non-specific symptoms |
| 2005 | Referral guidelines will help GPs identify patients with cancer | Full text unavailable |
| Alho, 2006 | Head and neck cancer in primary care: presenting symptoms and the effect of delayed diagnosis of cancer cases | No focus on non-specific symptoms |
| Abel, 2008 | Delays in referral and diagnosis for chronic hematologic malignancies: a literature review | No info on diagnostic reasoning |
| Jiwa, 2009 | Advice to consult a general medical practitioner in Western Australia: could it be cancer? | No info on diagnostic reasoning |
| Hamilton, 2010 | Cancer diagnosis in primary care | Wrong study design |
| Tate, 2010 | Are GPs under-investigating older patients presenting with symptoms of ovarian cancer? Observational study using General Practice Research Database | No focus on non-specific symptoms |
| Baughan, 2011 | Urgent suspected cancer referrals from general practice: audit of compliance with guidelines and referral outcomes | No focus on non-specific symptoms |
| Kovács, 2011 | [Delays in oncology care. Role of patients and of their family physicians] | No info on diagnostic reasoning |
| O'Neill, 2011 | Urgent referral for suspected cancer in Scotland | No focus on non-specific symptoms |
| Sprinks, 2011 | GPs slow to diagnose and refer teenage cancer patients | Wrong publication type |
| Van Hout, 2011 | Determinants of patient's and doctor's delay in diagnosis and treatment of colorectal cancer | No info on diagnostic reasoning |
| 2012 | Diagnosis delayed for certain cancers and certain patients | Wrong publication type |

**Table S6** (continued)

| **First author, date** | **Title** | **Reason for exclusion** |
| --- | --- | --- |
| 2012 | GPs take longer to refer women with cancer symptoms | Full text unavailable |
| Abel, 2012 | Referrals for suspected hematologic malignancy: a survey of primary care physicians | No focus on non-specific symptoms |
| Ahrensberg, 2012 | Presenting symptoms of children with cancer: a primary-care population-based study | No info on diagnostic reasoning |
| Johansen, 2012 | How does the thought of cancer arise in a general practice consultation? Interviews with GPs | No focus on non-specific symptoms |
| Treasure, 2013 | Symptoms and risk factors to identify people with suspected cancer in primary care | Wrong publication type |
| Davis, 2014 | Leukemia: an overview for primary care | Wrong study design |
| Jensen, 2014 | Quality deviations in cancer diagnosis: prevalence and time to diagnosis in general practice | No focus on non-specific symptoms |
| Neal, 2014 | Comparison of cancer diagnostic intervals before and after implementation of NICE guidelines: analysis of data from the UK General Practice Research Database | No info on diagnostic reasoning |
| 2015 | Tools will aid GPs in assessing people with possible cancer | Full text unavailable |
| Emery, 2015 | The challenges of early diagnosis of cancer in general practice | Wrong publication type |
| Green, 2015 | Cancer detection in primary care: insights from general practitioners | No focus on non-specific symptoms |
| Jones, 2015 | Recognition and referral of cancer symptoms in primary care | Wrong publication type |
| Mitchell, 2015 | The role of primary care in cancer diagnosis via emergency presentation: qualitative synthesis of significant event reports | No info on diagnostic reasoning |
| Neal, 2015 | The complexity and difficulty of diagnosing lung cancer: findings from a national primary-care study in Wales | No info on diagnostic reasoning |
| Rasmussen, 2015 | Specific and non-specific symptoms of colorectal cancer and contact to general practice | No info on diagnostic reasoning |
| 2016 | GPs need help diagnosing cancer | Wrong publication type |
| Losa Frías, 2016 | Early diagnosis of cancer in primary care | Wrong study design |
| 2017 | Missed and delayed colorectal cancer diagnoses may be attributed to process-of-care failures in primary care clinicians' workup | Wrong publication type |
| Koo, 2017 | Typical and atypical presenting symptoms of breast cancer and their associations with diagnostic intervals: Evidence from a national audit of cancer diagnosis | No info on diagnostic reasoning |
| Kostopoulou, 2017 | The role of physicians' first impressions in the diagnosis of possible cancers without alarm symptoms | No focus on non-specific symptoms |
| Sheringham, 2017 | Variations in GPs' decisions to investigate suspected lung cancer: a factorial experiment using multimedia vignettes | No focus on non-specific symptoms |
| Sirota, 2017 | Prevalence and alternative explanations influence cancer diagnosis: An experimental study with physicians | No focus on non-specific symptoms |

**Table S6** (continued)

| **First author, date** | **Title** | **Reason for exclusion** |
| --- | --- | --- |
| Aaronson, 2019 | Missed diagnosis of cancer in primary care: Insights from malpractice claims data | No info on diagnostic reasoning |
| Baun, 2019 | Ovarian cancer suspicion, urgent referral and time to diagnosis in Danish general practice: A population-based study | No info on diagnostic reasoning |
| Kostopoulou, 2019 | Referral decision making of general practitioners: A signal detection study | No info on diagnostic reasoning |
| Harris, 2020 | Primary care practitioners' diagnostic action when the patient may have cancer: an exploratory vignette study in 20 European countries | No focus on non-specific symptoms |
| Merriel, 2020 | Improving early cancer diagnosis in primary care | Wrong publication type |
| Nieminem, 2020 | Factors influencing patient and health care delays in oropharyngeal cancer | No focus on non-specific symptoms |
| Pearson, 2020 | Cross-sectional study using primary care and cancer registration data to investigate patients with cancer presenting with non-specific symptoms | No info on diagnostic reasoning |
| Ananth, 2021 | 67 Factors predicting lung cancer in urgent cancer referrals | Wrong publication type |
| Chapman, 2021 | Non-specific symptoms-based pathways for diagnosing less common cancers in primary care: A service evaluation | No info on diagnostic reasoning |
| Damhus, 2021 | Non-specific symptoms and signs of cancer: different organisations of a cancer patient pathway in Denmark | No info on diagnostic reasoning |
| Emery, 2021 | Approaches to diagnosing cancer earlier in general practice | Wrong publication type |
| Losa Frías, 2021 | How to suspect cancer in Primary Care | Wrong study design |
| Moodley, 2021 | Exploring primary care level provider interpretation and management of potential breast and cervical cancer signs and symptoms in South Africa | No focus on non-specific symptoms |
| Smith, 2021 | GPs’ use of gut feelings when assessing cancer risk: A qualitative study in UK primary care | No focus on non-specific symptoms |
| Virgilsen, 2021 | Alignment between the patient's cancer worry and the GP's cancer suspicion and the association with the interval between first symptom presentation and referral: a cross-sectional study in Denmark | No focus on non-specific symptoms |
| Virgilsen, 2021 | Patient's worry about cancer and the general practitioner's suspicion of cancer or serious illness: A population-based study in Denmark | No focus on non-specific symptoms |
| Arreskov, 2022 | General practitioner responses to concerns in chronic care consultations for patients with a history of cancer | No info on diagnostic reasoning |
| Hardy, 2022 | Role of primary care physician factors on diagnostic testing and referral decisions for symptoms of possible cancer: A systematic review | No focus on non-specific symptoms |

**Table S6** (continued)

| **First author, date** | **Title** | **Reason for exclusion** |
| --- | --- | --- |
| Jensen, 2022 | The pathway and characteristics of patients with non-specific symptoms of cancer: a systematic review | No info on diagnostic reasoning |
| Rampes, 2022 | Early diagnosis of symptomatic ovarian cancer in primary care in the UK: opportunities and challenges | Wrong study design |
| Saab, 2022 | A systematic review of interventions to recognise, refer and diagnose patients with lung cancer symptoms | No focus on non-specific symptoms |
| Tzanis, 2022 | What factors empower general practitioners for early cancer diagnosis? A 20-country European Delphi Study | No focus on non-specific symptoms |
| Black, 2023 | What causes delays in diagnosing blood cancers? A rapid review of the evidence | Wrong study design |
| Devi, 2023 | Identification of barriers at the primary care provider level to improve inflammatory breast cancer diagnosis and management | No info on diagnostic reasoning |
| Gbenjo, 2023 | Leukemia: What primary care physicians need to know | Wrong study design |
| Lamprell, 2023 | People with early-onset colorectal cancer describe primary care barriers to timely diagnosis: a mixed-methods study of web-based patient reports in the United Kingdom, Australia and New Zealand | No info on diagnostic reasoning |
| Lauridsen, 2023 | Exploring GPs’ assessments of their patients’ cancer diagnostic processes: a questionnaire study | No info on diagnostic reasoning |
| O'Neill, 2023 | Referral challenges for early-onset colorectal cancer: a qualitative study in UK primary care | No focus on non-specific symptoms |
| Rao, 2023 | Recognition, diagnostic practices, and cancer outcomes among patients with unintentional weight loss (UWL) in primary care | No relation with cancer |
| Yao, 2023 | Gut feeling for the diagnosis of cancer in general practice: A diagnostic accuracy review | No focus on non-specific symptoms |
| Damhus, 2024 | Diagnostic flow for all patients referred with non-specific symptoms of cancer to a diagnostic centre in Denmark: A descriptive study | No info on diagnostic reasoning |
| Koskela, 2024 | What would primary care practitioners do differently after a delayed cancer diagnosis? Learning lessons from their experiences | No focus on non-specific symptoms |
