## Supplemental Table 7 for "Attributing non-specific symptoms to cancer in general practice: a scoping review"

### S3 Table. Search strategy for PsycInfo via Ovid (26/03/2024)

| **Searches** | **Query** | **Results** |
| --- | --- | --- |
| 1 | exp *Neoplasms/ | 56960 |
| 2 | (adenocarcinoma* or blastoma* or cancer* or carcinoma* or glioblastoma* or gonadoblastoma* or hepatoblastoma* or hodgkin* or leuk?emia* or lymphoma* or malignan* or medulloblastoma* or melanoma* or myeloma* or neoplas* or nephroblastoma* or neuroblastoma* or nonhodgkin* or "non-hodgkin*" or onco* or pancreatoblastoma* or retinoblastoma* or sarcoma* or tumo*).ti. | 57217 |
| 3 | 1 or 2 | 67850 |
| 4 | exp General Practitioners/ | 6525 |
| 5 | "generalist*".ab,ti. | 2507 |
| 6 | exp Family Medicine/ | 1366 |
| 7 | exp Family Physicians/ | 1661 |
| 8 | ((family or general) adj (doctor* or medicine or physician* or practi*)).ti,ab. | 22570 |
| 9 | (GP or GPs).ti,ab. | 9128 |
| 10 | exp primary health care/ | 22211 |
| 11 | (primary adj2 care).ab,ti. | 40133 |
| 12 | 4 or 5 or 6 or 7 or 8 or 9 or 10 or 11 | 67533 |
| 13 | "clinical reasoning".ab,ti. | 1465 |
| 14 | "causal attribution".ab,ti. | 1197 |
| 15 | "assess*".ab,ti. | 910030 |
| 16 | "clinical decision-making".ab,ti. | 3887 |
| 17 | "attribut*".ab,ti. | 142855 |
| 18 | "first impression".ab,ti. | 387 |
| 19 | perception.ab,ti. | 187637 |
| 20 | "recogni*".ab,ti. | 236237 |
| 21 | (suspicion* or suspicious or suspect*).ab,ti. | 24592 |
| 22 | "apprais*".ab,ti. | 37964 |
| 23 | (think or thinks or thought or thinking).ab,ti. | 243256 |
| 24 | (refer* not (reference* or referencing)).ab,ti. | 145483 |
| 25 | referral*.ti,ab. | 32387 |
| 26 | ((refer adj2 patient) or (refers adj2 patient) or (refer adj2 patients) or (refers adj2 patients)).ti,ab. | 885 |
| 27 | 13 or 14 or 15 or 16 or 17 or 18 or 19 or 20 or 21 or 22 or 23 or 24 or 25 or 26 | 1678456 |
| 28 | 3 and 12 and 27 | 932 |
