## Supplemental Table 4 for "Attributing non-specific symptoms to cancer in general practice: a scoping review"

### S4 Table. Search strategy for Embase (26/03/2024)

| **No.** | **Query** | **Results** |
| --- | --- | --- |
| #1 | neoplasm'/exp/mj | 4,706,279 |
| #2 | adenocarcinoma*:ti OR blastoma*:ti OR cancer*:ti OR carcinoma*:ti OR glioblastoma*:ti OR gonadoblastoma*:ti OR hepatoblastoma*:ti OR hodgkin*:ti OR leuk?emia*:ti OR lymphoma*:ti OR malignan*:ti OR medulloblastoma*:ti OR melanoma*:ti OR myeloma*:ti OR neoplas*:ti OR nephroblastoma*:ti OR neuroblastoma*:ti OR nonhodgkin*:ti OR 'non-hodgkin*':ti OR onco*:ti OR pancreatoblastoma*:ti OR retinoblastoma*:ti OR sarcoma*:ti OR tumo*:ti | 3,978,722 |
| #3 | #1 OR #2 | 5,393,792 |
| #4 | 'general practitioner'/exp | 123,155 |
| #5 | 'general practice'/exp | 93,422 |
| #6 | generalist*:ti,ab | 12,78 |
| #7 | ((family OR general) NEXT/1 (doctor* OR medicine OR physician* OR practi*)):ti,ab | 182,4 |
| #8 | gp:ti,ab OR gps:ti,ab | 105,338 |
| #9 | 'primary health care'/exp | 215,494 |
| #10 | (primary NEAR/2 care):ti,ab | 233,278 |
| #11 | 'primary care':ti,ab | 200331 |
| #12 | #4 OR #5 OR #6 OR #7 OR #8 OR #9 OR #10 OR #11 | 573490 |
| #13 | 'clinical reasoning'/exp | 2827 |
| #14 | 'clinical reasoning':ti,ab | 5805 |
| #15 | 'patient assessment'/exp | 34475 |
| #16 | assess*:ti,ab | 5663307 |
| #17 | 'causal attribution'/exp | 10011 |
| #18 | 'causal attribution':ti,ab | 559 |
| #19 | 'clinical decision making':ti,ab | 39271 |
| #20 | attribut*:ti,ab | 585050 |
| #21 | 'first impression':ti,ab | 707 |
| #22 | 'perception'/exp | 543242 |
| #23 | perception:ti,ab | 272583 |
| #24 | 'recognition'/exp | 38655 |
| #25 | recogni*:ti,ab | 1202323 |
| #26 | suspicion*:ti,ab OR suspicious:ti,ab OR suspect*:ti,ab | 559094 |
| #27 | apprais*:ti,ab | 91256 |
| #28 | think:ti,ab OR thinks:ti,ab OR thought:ti,ab OR thinking:ti,ab | 501618 |
| #29 | 'patient referral'/exp | 166694 |
| #30 | refer*:ti,ab NOT (reference*:ti,ab OR referencing:ti,ab) | 740698 |
| #31 | referral*:ti,ab | 248176 |
| #32 | ((refer NEAR/2 patient):ti,ab) OR ((refers NEAR/2 patient):ti,ab) OR ((refer NEAR/2 patients):ti,ab) OR ((refers NEAR/2 patients):ti,ab) | 8534 |
| #33 | #13 OR #14 OR #15 OR #16 OR #17 OR #18 OR #19 OR #20 OR #21 OR #22 OR #23 OR #24 OR #25 OR #26 OR #27 OR #28 OR #29 OR #30 OR #31 OR #32 | 8998742 |
| #34 | #3 AND #12 AND #33 | 17,391 |
| #35 | #34 AND ('Article'/it OR 'Review'/it) | 10,425 |
| #36 | #35 AND 'case report'/de | 799 |
| #37 | #35 NOT #36 | 9,626 |
