## Supplemental Table 2 for "Attributing non-specific symptoms to cancer in general practice: a scoping review"

### S2 Table. Search strategy for Medline via PubMed (26/03/2024)

| **Search** | **Query** | **Filters** | **Results** |
| --- | --- | --- | --- |
| #1 | "Neoplasms"[Majr] |  | 3.513.931 |
| #2 | adenocarcinoma*[TI] OR blastoma*[TI] OR cancer*[TI] OR carcinoma*[TI] OR glioblastoma*[TI] OR gonadoblastoma*[TI] OR hepatoblastoma*[TI] OR hodgkin*[TI] OR leukaemi*[TI] OR leukemi*[TI] OR lymphoma*[TI] OR malignan*[TI] OR medulloblastoma*[TI] OR melanoma*[TI] OR myeloma*[TI] OR neoplas*[TI] OR nephroblastoma*[TI] OR neuroblastoma*[TI] OR nonhodgkin*[TI] OR "non-hodgkin*"[TI] OR onco*[TI] OR pancreatoblastoma*[TI] OR retinoblastoma*[TI] OR sarcoma*[TI] OR tumour*[TI] OR tumor*[TI] |  | 3.073.143 |
| #3 | #1 OR #2 |  | 4.126.117 |
| #4 | "Family Practice"[Mesh] |  | 67.446 |
| #5 | "General Practice"[Mesh] |  | 79.098 |
| #6 | "General Practitioners"[Mesh] |  | 11.122 |
| #7 | "Family doctor*"[TIAB] OR "Family medicine"[TIAB] OR "Family physician*"[TIAB] OR "Family practice*"[TIAB] OR "Family practitioner*"[TIAB] OR "General doctor*"[TIAB] OR "General medicine"[TIAB] OR "General physician*"[TIAB] OR "General practice*"[TIAB] OR "General practitioner*"[TIAB] |  | 139.612 |
| #8 | GP[TIAB] OR GPs[TIAB] |  | 74.020 |
| #9 | "Primary Health Care"[Mesh] |  | 195.993 |
| #10 | "primary care"[TIAB] OR "primary care"[TIAB:~2] |  | 188.732 |
| #11 | #4 OR #5 OR #6 OR #7 OR #8 OR #9 OR #10 |  | 485.007 |
| #12 | "Clinical Decision-Making"[Mesh] |  | 16.735 |
| #13 | "Clinical Decision-Making"[TIAB] |  | 29.195 |
| #14 | "Clinical Reasoning"[Mesh] |  | 1.891 |
| #15 | "clinical reasoning"[TIAB] |  | 4.869 |
| #16 | "Symptom Assessment"[Mesh] |  | 7.113 |
| #17 | assess*[TIAB] |  | 4.007.877 |
| #18 | attribut*[TIAB] |  | 485.650 |
| #19 | apprais*[TIAB] |  | 75.203 |
| #20 | "first impression"[TIAB] |  | 526 |
| #21 | perception[TIAB] |  | 233.285 |
| #22 | recogni*[TIAB] |  | 945.838 |
| #23 | suspicion*[TIAB] OR suspicious[TIAB] OR suspect*[TIAB] |  | 355.641 |
| #24 | Think[TIAB] OR Thinks[TIAB] OR Thought[TIAB] OR Thinking[TIAB] |  | 370.511 |
| #25 | reasoning[TIAB] |  | 30.755 |
| #26 | "Referral and Consultation"[Mesh] |  | 88.014 |
| #27 | refer*[TIAB] NOT (reference*[TIAB] OR referencing[TIAB]) |  | 476.817 |
| #28 | Referral*[TIAB] |  | 150.476 |
| #29 | "refer patient"[TIAB:~2] OR "refers patient"[TIAB:~2] OR "refer patients"[TIAB:~2] OR "refers patients"[TIAB:~2] |  | 5.813 |
| #30 | #12 OR #13 OR #14 OR #15 OR #16 OR #17 OR #18 OR #19 OR #20 OR #21 OR #22 OR #23 OR #24 OR #25 OR #26 OR #27 OR #28 OR #29 |  | 6.375.746 |
| #31 | #3 AND #11 AND #30 |  | 12.491 |
| #32 | #3 AND #11 AND #30 | Case Reports | 675 |
| #33 | #31 NOT #32 |  | 11.816 |
