## Supplemental Table 5 for "Attributing non-specific symptoms to cancer in general practice: a scoping review"

### S5 Table. Search strategy for Open Access Theses and Dissertation (11/07/2024)

| **Num** | **Search** | **Hits** |
| --- | --- | --- |
| 8 | abstract:("general practitioner") AND abstract:(cancer) AND abstract:("diagnosis") AND pub_dt:[2022-01-01T00:00:00Z TO *] | 4 |
| 7 | title:("general practitice") AND ("cancer diagnosis") | 3 |
| 6 | abstract:(GP) AND abstract:("cancer diagnosis") AND pub_dt:[2022-01-01T00:00:00Z TO *] | 2 |
| 5 | abstract:(diangostic AND reasoning) AND abstract:(cancer) AND abstract:("general practice") | 1 |
| 4 | abstract:("think of cancer") AND abstract:(general OR practice) AND pub_dt:[2022-01-01T00:00:00Z TO *] | 0 |
| 3 | abstract:("clinical reasoning") AND abstract:(general OR practice) AND abstract:("cancer") | 4 |
| 2 | abstract:("think of cancer") AND abstract:(general OR practice) AND pub_dt:[2022-01-01T00:00:00Z TO *] | 0 |
| 1 | abstract:(diagnostic AND reasoning) AND abstract:(cancer) AND abstract:("general practice") | 1 |
